# Comparison of total and neutralizing SARS-CoV-2 spike antibodies against omicron and other variants in paired samples after two or three doses of mRNA vaccine

**DOI:** 10.1101/2022.01.26.22269819

**Authors:** Amanda K. Debes, Shaoming Xiao, Emily R. Egbert, Patrizio Caturegli, Ioannis Sitaras, Andrew Pekosz, Aaron M. Milstone

**Affiliations:** Johns Hopkins University Bloomberg School of Public Health, Baltimore, MD, USA; Johns Hopkins University School of Medicine, Baltimore, MD, USA

## Abstract

Recognizing that anti-SARS-CoV-2 antibody levels wane over time following the 2-dose SARS-CoV-2 mRNA series, the FDA approved a booster dose for people greater than 12 years old. Limited data exist on whether a booster dose of the mRNA vaccine results in greater antibody protection than the primary series. We examined total and neutralizing antibodies to the spike protein of SARS-CoV-2, and neutralizing antibodies against Washington-1 (WA-1) and variants of concern (VOC) including Beta, Delta and Omicron in a longitudinal cohort. Healthcare workers (HWs) were included in the analysis if serum was collected 1) within 14-44 days post-dose2 of an mRNA SARS-CoV-2 vaccine (Timepoint 1, TP1), or 2) at least 8 months post-dose2 (Timepoint 2, TP2), or 3) within 14-44 days following mRNA booster (Timepoint 3, TP3). HWs with prior covid-positive PCR were excluded. We found that there is little to no neutralizing capability following a 2-dose mRNA vaccine series against the omicron variant, and neutralizing capacity to any variant strain tested has been lost by 8-months post two-dose vaccination series. However, the mRNA booster series eliminates the immune escape observed by the omicron variant with the two-dose series. Neutralizing titers were significantly higher for all variants post-boost compared to the titers post two-dose series. The longitudinal nature of our cohort facilitated the analysis of <u>paired</u> samples pre and post boost, showing a greater than 15-fold increase in neutralization against omicron post-boost in these paired samples. An mRNA booster dose provides greater quantity and quality of antibodies compared to a two-dose regimen and is critical to provide any protection against the omicron variant.

## Introduction

SARS-CoV-2 antibody levels wane following two-dose mRNA vaccination and infection.^1^ mRNA booster doses are available and protect against hospitalization and death, but booster uptake remains low.^2^ Our objective was to compare total and neutralizing SARS-CoV-2 spike antibodies against against Washington-1 (WA-1) and variants of concern (VOC) in a longitudinal cohort.

## Methods

Healthcare workers (HWs) were consented into a seroprevalence cohort beginning June 2020 and followed through November 2021.^1^ HWs provided serum samples longitudinally and were included in this analysis if serum was collected 1) within 14-44 days post-dose2 of an mRNA SARS-CoV-2 vaccine (Timepoint 1, TP1), or 2) at least 8 months post-dose2 (Timepoint 2, TP2), or 3) within 14-44 days following mRNA booster (Timepoint 3, TP3). HWs with prior covid-positive PCR were excluded. To determine if the increase in magnitude of antibody response to the mRNA vaccine strain, as measured by enzyme-linked immunosorbent assay (ELISA) [Euroimmun], led to an increased recognition of VOC, neutralizing antibody titer (NT) assays were performed against the vaccine strain (WA-1) and the Beta, Delta and Omicron variants as previously described.^1,3^ For NT assays, 45 HWs were selected, prioritizing paired samples from TP1 and TP3, and others were selected at random.

Wilcoxon rank sum test was used for unpaired analyses, and Wilcoxon signed rank test and Friedman test for paired analyseis. The Johns Hopkins University Institutional Review Board approved this study. Analyses were performed in R, version 4.1.2.

## Results

Of 3032 HWs originally enrolled in the longitudinal cohort, 1353 contributed serum to at least one of the 3 groups: 507 within in TP1, 879 in TP2, and 273 in TP3. Of these 1353 participants, 81% were women, 96% were Non-Hispanic/Latino, and 81% were White. The median (IQR) age of participants was 41.8 (33.8 - 53.3) years.

High levels of antibodies in TP1 waned to lower levels in TP2 and then boosted to much higher levels in TP3. Of the TP3 samples tested, 94% demonstrated spike IgG assay saturation compared to 59% in TP1 (Figure 1). Spike IgG measurements correlated with NT against WA-1. TP1 samples had lower NT activity across VOC compared to WA-1 (Figure 2). By TP2, there was little NT to Beta and Delta, and none to omicron (titer<20).^4^ At TP3, NT activity against all viruses was boosted and the fold reductions between WA-1 and VOC were less than those observed in TP1 among paired samples.

**Figure 1:**
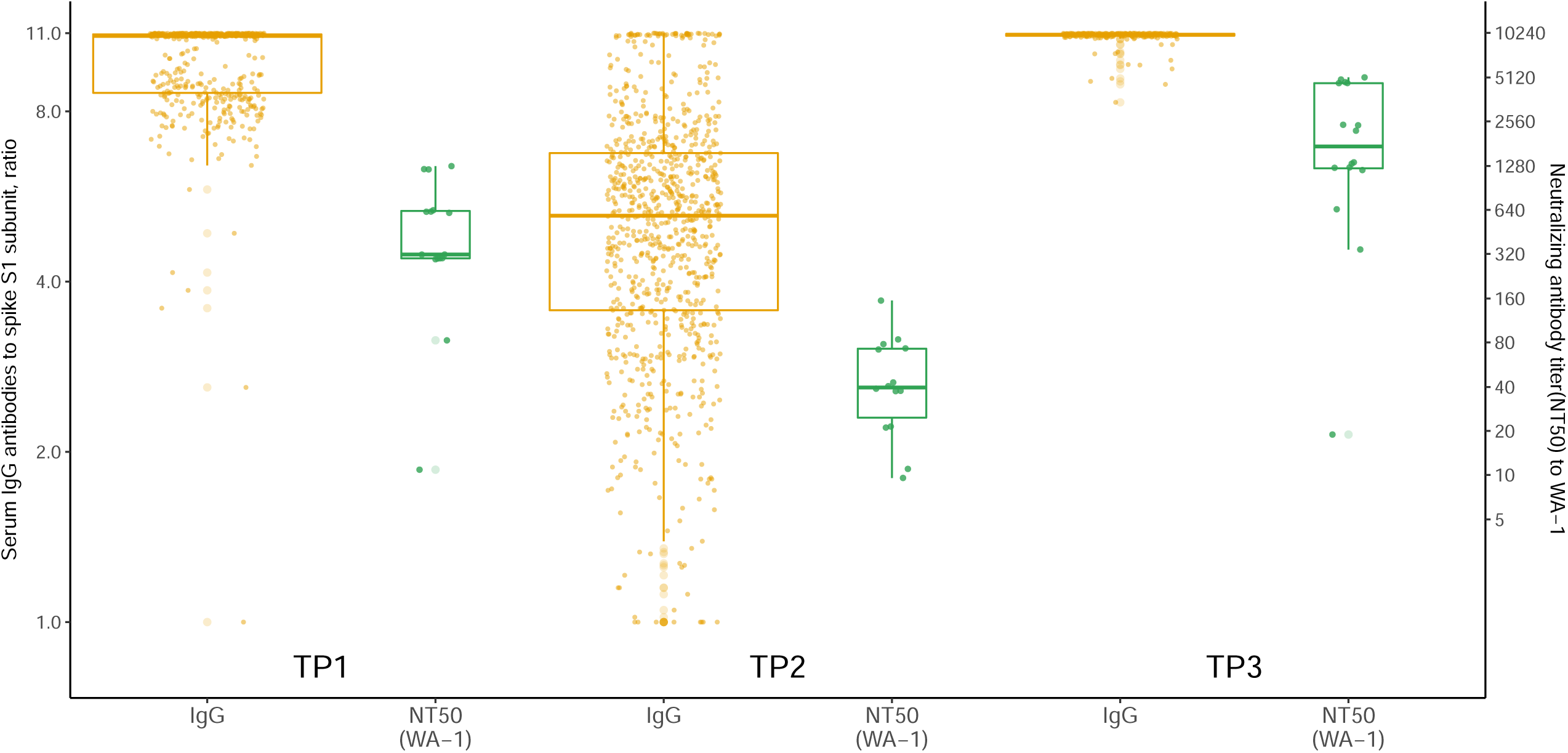
Spike IgG serum antibodies and live-virus neutralizing antibody titers (NT) against the vaccine strain (WA-1). Data shown at threetime points: within 14-44 days post-dose2 (Timepoint 1), at least 8 months post-dose2 (Timepoint 2), and within 14-44 post-booster (Timepoint 3). IgG antibody measurements estimated optical density ratios with a lower threshold of 1.23 and upper threshold of 11.00 based on assay saturation. NTs were reported using NT50, and a positive threshold was defined as NT≥20.^4^ Timepoint 1: 396(78%) females, 484(95%) non-Hispanic/Latino, 383(76%) got Pfizer for primary dose, 395 (78%) whites, median age(IQR): 39.9(32.4, 51.9) Timepoint 2: 739(84%) females, 845(96%) non-Hispanic/Latino, 657(75%) got Pfizer for primary dose, 723 (82%) whites, median age(IQR): 43.0(35.1, 53.5) Timepoint 3: 215(79%) females, 263(96%) non-Hispanic/Latino, 225(82%) got Pfizer for primary dose, 238 (87%) whites, median age(IQR): 44.9(34.3, 55.6). NT were performed at Timepoint 1 (n=15), Timepoint 2 (n=14), Timepoint 3 (n=16).

**Figure 2:**
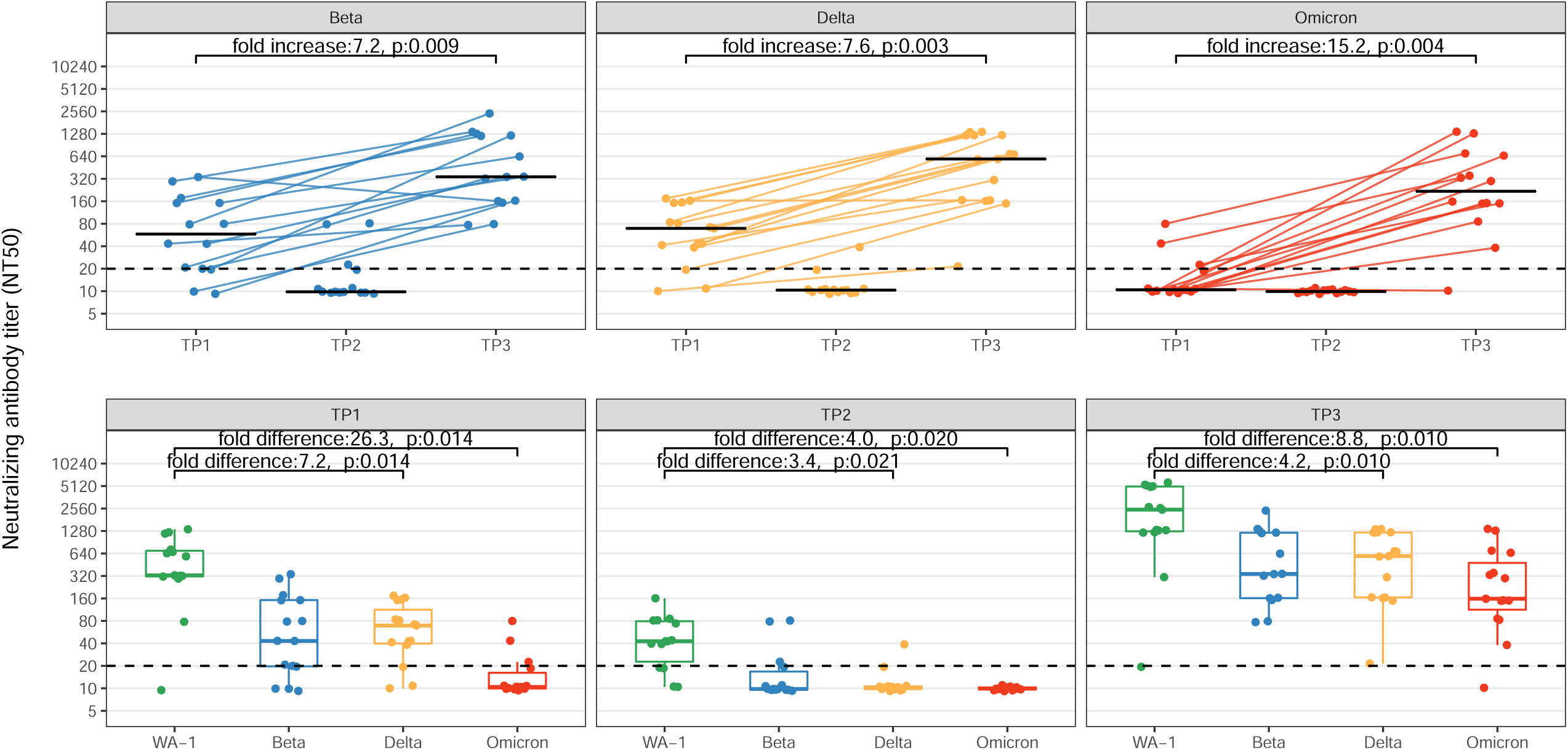
Comparison of neutralizing antibody titers (NT) to SARS-CoV-2 vaccine strain (WA-1), Beta, Delta, and Omicron VOC from healthcare workers with paired serum samples in a longitudinal cohort. Data shown at three timepoints: within 14-44 days post-dose-2 (Timepoint 1), at least 8 months post-dose-2 (Timepoint 2), and within 14-44 post-booster (Timepoint 3). Top panel shows NT titer for each variant across the three timepoints with connecting lines illustrating 15 paired samples in Timepoints 1 and 3. Bottom panel shows NT at each timepoint for each VOC. Fold change (increase/difference) represents geometric median fold change. P-values have been corrected for multiple comparisons using Bonferroni methods.

## Discussion

After two-dose vaccination, HWs developed high spike IgG and NT to vaccine strains but significantly lower NT against VOC was evident. By 8-month post-dose-2, all antibody measures had waned and NT against VOC had dropped to nearly undetectable levels. Boosting proved critical, as NT were significantly higher for all VOC compared to post-dose2 titers, including a >15 fold increase in neutralization against omicron in paired samples.

Limitations include that paired serum was only available post-dose2 and post-boost, and nABs titers were only performed on a small subset. The corresponding antibody trends between serum IgG and NT supports that despite only testing a subset of the participants for virus neutralization, all participants likely had robust broad NT post-boost, findings supported by recently published pre and post-boost studies.^5^

This study demonstrates that in paired samples an mRNA vaccine booster produces greater quantity and function of spike antibodies and NT as compared to primary SARS-CoV-2 mRNA immunization and was necessary to restore measurable NT to VOC. The booster dose eliminated the immune escape observed by Omicron following two-dose mRNA immunization. The variable NT to mRNA booster and whether breakthrough omicron infection boosts NT must be investigated to understand durable immunity against future VOC.

## Data Availability

All data produced in the present study are available upon reasonable request to the authors.

## Acknowledgements

The authors would like to thank members of the Johns Hopkins Hospital Clinical Immunology Laboratory, Danielle Koontz and Ani Voskertchian of the Johns Hopkins Division of Pediatric Infectious Diseases, Elizabeth Colantuoni from the Johns Hopkins Bloomberg School of Public Health, and Avinash Gadala from the Johns Hopkins Health System. Research reported in this publication was supported in part by the National Institute of Allergy and Infectious Diseases of the National Institutes of Health (NIH) under award number K24AI141580 (A.M.), the Johns Hopkins Center of Excellence in Influenza Research and Surveillance (NIH/NIAID N2772201400007C AP) and the generosity of the collective community of donors to the Johns Hopkins University School of Medicine and the Johns Hopkins Health System for Covid-19 research.

## Conflict of Interest Disclosure

A.M. reports grant support from Merck for work unrelated to this study. Other authors report no conflicts.

## Notes

### Competing Interest Statement

The authors have declared no competing interest.

### Author Declarations

The Johns Hopkins University Institutional Review Board gave ethical approval for this study.

### Summary of Updates

There was a typo in one of the co-authors last names that needed to be fixed for the abstract (it is correct in the pdf).

